# Proportion of new genital human papillomavirus detections attributable to latent infections: Implications for cervical cancer screening

**DOI:** 10.1101/2021.05.12.21256547

**Authors:** Talía Malagón, Aaron MacCosham, Ann N. Burchell, Mariam El-Zein, Pierre-Paul Tellier, François Coutlée, Eduardo L. Franco, the HITCH Study Group

## Abstract

**Background:** Infections with human papillomaviruses (HPV) may enter into a latent state in epithelial basal cells, and eventually become reactivated following loss of immune control. It is unclear what proportion of incident detections of HPV are due to reactivation of previous latent infections versus new transmissions.

**Methods:** The HITCH cohort study prospectively followed young newly-formed heterosexual partners recruited between 2005-2011 in Montréal, Canada. We calculated the fraction of incident HPV detections non-attributable to sexual transmission risk factors with a Bayesian Markov state transition model. Results are the median (2·5-95·5^th^ percentiles) of the estimated posterior distribution.

**Findings:** 544 type-specific incident HPV detection events occurred in 849 participants; 32·5% of all incident HPV detections occurred in participants whose HITCH partners were negative for that HPV type and who did not report having sex with anyone else over follow-up. We estimate that 42·7% (38·4-47·2%) of all incident HPV detections in this population might be attributable to reactivation of latent infections, not transmission.

**Interpretation:** A positive HPV test result in many cases may be a reactivated past infection, rather than a new infection from recent sexual behaviors or partner infidelity. The potential for reactivation of latent infections in previously HPV-negative women should be considered in the context of cervical cancer screening.

**Funding:** Canadian Institutes of Health Research, National Institutes of Health, Merck-Frosst Canada Ltd, Merck & Co Ltd, Fonds de la Recherche en Santé du Québec.

**Research in Context:** *Evidence before this study:* Previous studies assessing the proportion of HPV infections attributable to reactivation in women have been conducted in individual-based studies. Determining this estimate using a couple-based study design could account for the partner’s HPV status and rule out sexual transmission. Authors from this current study recently published a systematic review that aimed to identify all published couple-based studies measuring HPV transmission. They searched MEDLINE, EMBASE, Scopus, and Cochrane Library from database inception to December 1, 2019, with no language restrictions using the keywords and MeSH terms “HPV,” “papillomavirus infections,” “papillomaviridae,” “transmission,” “heterosexuality,” “couples,” and “sexual partners”. Studies were included if the study population was heterosexual couples, genital samples were collected from each partner, and HPV transmission rates were reported. The search yielded 834 records, of which seven couple-based studies were eligible to be included in the systematic review. None of the identified studies measured the proportion of HPV infections attributable to reactivation.

*Added value of this study:* This study presents the first analysis assessing reactivation of HPV infections using a couple-based study design. We recruited young heterosexual couples and collected genital HPV data from both partners, allowing us to control for the sexual partner’s HPV status. We estimate that 57% of the newly detected incident HPV infections in women could be attributed to sexual transmission while the remaining 43% is most likely due to reactivation of latent infections.

*Implications of all the available evidence:* In the context of cervical cancer screening, our findings suggest that women who have previously tested HPV negative may not remain HPV-negative, even with no new sexual partners, due to reactivation of a latent infection. This underscores the importance for HPV-negative women to undergo multiple screenings in their lifetime. In addition, the sizeable proportion of newly detected HPV infections attributable to reactivation suggests that a positive HPV test is not necessarily due to recent sex or partner infidelity, which may help de-stigmatize a positive HPV test result.

## Background

Human papillomaviruses (HPV) are sexually transmitted infections causing anogenital and oropharyngeal cancers.^1^ Biological evidence suggests that, following immune control of the initial infection, many HPV infections may enter into a latent state in epithelial basal cells.^2^ These latent infections may persist for years, and are generally undetectable with HPV DNA tests as they produce no or very low viral copy numbers. Latent infections may later become reactivated and re-detectable following loss of immune control, inflammation, or immunosuppression.^2,3^ Evidence suggests that HPV redetections have a comparable risk of oncogenic progression as first detections.^4^ Incident HPV detections in older women, which are more likely to be redetections, appear to carry comparable risks of progression to high-grade cervical lesions as incident detections younger women.^5^ As more countries adopt primary HPV testing for cervical cancer screening, the potential for reactivation of latent infections in women who have previously tested negative for HPV should be considered.

Given the difficulty in ruling out sexual transmission in human studies, it is yet unclear what proportion of incident detections of HPV are due to recent transmission from a sexual partner, or to a reactivated latent infection. While many studies have found that incident HPV detections occur in women who report no new sexual partners, they generally cannot rule out transmission from a current sexual partner and have no data on partner HPV status.^6,7^ We used partner data from the HPV Infection and Transmission among Couples through Heterosexual activity (HITCH) cohort study to estimate the proportion of incident genital HPV detections attributable to transmission as opposed to reactivation.

## Methods

### Study design & setting

The full study protocol, procedures, and data collection instruments for the HITCH cohort study have been published previously.^8-11^ Briefly, the study enrolled young female university and college students aged 18-24 years old and their male sex partners ≥18 years old in Montréal, Canada between 2005-2011. Participants had to be in a new (≤6 months) sexual partnership. Participants were followed-up over 2 years for women (visits at 0, 4, 8, 12, 18, and 24 months), and 4 months for men (visits at 0 and 4 months). All participants provided written informed consent. The ethical review committees of McGill University, Concordia University, and the Centre Hospitalier de l’Université de Montreal approved the study.

At each clinic visit, participants completed a self-administered questionnaire and provided genital samples for HPV testing. Women self-collected vaginal samples using a Dacron swab. The nurse collected male epithelial cells from the penis and scrotum in separate sample containers using gentle exfoliation with ultra-fine emery paper followed by swabbing with a Dacron swab. Samples were tested for HPV DNA using the Linear Array HPV genotyping assay (LA-HPV) (Roche Molecular Systems, Alameda, CA, USA),^12^ which detects 36 different HPV genotypes.

In this analysis, we used the data from the follow-up visit questionnaires, where participants reported if they had engaged in sex with anyone else other than their HITCH partner since the previous visit. Sex was defined as any sexual activity (hand, oral, vaginal, or anal).

### Statistical analysis

We restricted this analysis to the 447 women and 402 men in HITCH with at least two study visits with valid genital HPV samples. We furthermore restricted the analysis to the follow-up intervals between two study visits where a participant’s partner’s HPV status was known at the start of the interval (the first of the two visits). Demographic and clinical details on this subsample of the HITCH cohort have been previously described elsewhere.^13^

The outcome of interest was incident type-specific HPV detection, defined as a new detection of an HPV type in a participant who was previously negative for that type. We considered that risk factors for sexual HPV transmission were 1) having a HITCH partner positive for that HPV type or 2) having sex (hand, oral, vaginal, or anal) with other people during follow-up. These variables were treated as time-varying risk factors for participants followed-up over multiple intervals. We calculated the proportion of incident type-specific HPV detections attributable to transmission using the formula for the population attributable fraction (PAF):^14^

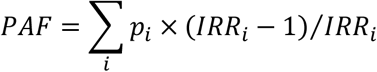

Where *i* is the level of a categorical risk factor for HPV transmission, *p*_*i*_ is the proportion of cases occurring in that category of the risk factor, and *IRR*_*i*_ is the HPV type-specific incidence rate ratio (IRR) associated with that category of the risk factor. We assumed, similarly to others,^6^ that all HPV detections non-attributable to transmission (1-PAF) were those potentially attributable to latent reactivation.

While HPV status is known for all partners at the start of each interval between visits, the partner HPV status at the end of the interval between visits is unknown for 48% of intervals either by design (men were followed over fewer visits than women) or due to partners being lost to follow-up. In 28% of cases, loss to follow-up was due to the relationship reportedly ending during the interval, resulting in one of the partners dropping out of the study, or coming for testing on a different day than their ex-partner, often weeks or months apart. Since transmission is less likely for these partnerships, and because we wanted to attribute incident HPV detections to sexual transmission only when there was evidence that a partner was infected with the same HPV type either previously or concurrently, we treated partner HPV status as missing for these visits and combined missing values with the ‘No’ category in the analysis. One participant (0·1%) did not answer the question relating to whether they had other sexual partners in the interval; we assigned this participant to the ‘No’ category for this variable.

We estimated type-specific HPV incidence rates, IRRs, and PAFs using a Bayesian Markov multistate model. The model estimates HPV incidence rates adjusting for interval-censoring and clearance. The unit of analysis was the HPV type, so participants provided multiple observations with each HPV type; the model accounts for multiple observations per participant with participant-specific random effects for estimation of HPV incidence rates. Details of the model have been described previously.^13^ The model was implemented using WinBUGS version 1.4.3 and the R2WinBUGS package in R version 3.6.3.^15,16^ Code for the current analysis is provided as supplementary material. The posterior distribution was estimated based on three independent Markov chains run for 50,000 iterations, with a burn-in of 20,000 iterations and a thinning factor of 5. For each estimate, we present the median of the posterior distribution, and its 2·5^th^ and 97·5^th^ percentiles as the 95% posterior probability interval (95%PI).

## Results

There were 544 type-specific incident HPV detection events during this follow-up, 314 in women and 230 in men (Table 1). The median follow-up time was 6·4 months per participant (interquartile range 4·6-9·7), 8.6 months (interquartile range 6·21-11·56) for women and 5·2 months (interquartile range 4·14-6·51) for men.

**Table 1.** Incidence and fraction of all type-specific HPV detections attributable and non-attributable to sexual transmission risk factors.

| Sex | Partner same type HPV positive (start of interval) | Partner same type HPV positive (end of interval) | Other sex partners | Incident HPV detections n (%) | Incidence rate (/100 person-months) | IRR | (95% PI) |
| --- | --- | --- | --- | --- | --- | --- | --- |
| Overall | Yes | .. | .. | 166 (30.5%) | 3.9 | 37.0 | (28.9-47.3) |
|  | No | Yes | .. | 72 (13.2%) | 6.6 | 62.7 | (44.9-87.6) |
|  | No | No/Missing | Yes | 129 (23.7%) | 0.3 | 2.7 | (2.1-3.6) |
|  | No | No/Missing | No<sup>a</sup> | 177 (32.5%) | 0.1 | Ref | .. |
| Women | Yes | .. | .. | 106 (33.8%) | 3.6 | 41.3 | (30.1-57.2) |
|  | No | Yes | .. | 36 (11.5%) | 5.5 | 63.1 | (39.9-100.0) |
|  | No | No/Missing | Yes | 80 (25.5%) | 0.3 | 3.0 | (2.1-4.3) |
|  | No | No/Missing | No | 92 (29.3%) | 0.1 | Ref | .. |
| Men | Yes | .. | .. | 60 (26.1%) | 4.5 | 32.1 | (21.9-47.6) |
|  | No | Yes | .. | 36 (15.7%) | 8.5 | 61.1 | (37.4-100.5) |
|  | No | No/Missing | Yes | 49 (21.3%) | 0.4 | 2.7 | (1.7-4.2) |
|  | No | No/Missing | No<sup>a</sup> | 85 (37.0%) | 0.1 | Ref | .. |

IRR=Incidence rate ratio; PI=prediction interval; Ref=Reference level
<sup>a</sup>One participant with missing data for other sex partners was included in this category

Having a HITCH partner positive for a given HPV type was a strong risk factor for incident HPV detection with that type. The incidence rate of type-specific HPV detection was 37·0 (95%PI: 28·9-47·3) times higher in participants whose partner was positive for that HPV type at the start of the interval, and 62·7 (95%PI: 44·9-87·6) times higher in participants whose partner was positive for that HPV type at the end of the interval, than in participants whose partner had no evidence of being positive for that HPV type and who reported no other sex partners in the interval. Having other sex partners was also a risk factor for incident HPV detection. Participants whose HITCH partner was negative at the start and negative/missing at end of the interval for that HPV type, but who reported having other sex partners had a 2·7 (95%PI: 2·1-3·6) times higher incidence rate of type-specific HPV detection than participants who did not report other sex partners in the interval. While the incidence rate of type-specific HPV detection was very low in participants whose HITCH partner was negative for that HPV type and who did not report other sex partners (0·1/10 person-years), 32·5% of all incident HPV detections occurred in this category of participants.

Using the PAF formula, we estimated that 29·6% (95%PI: 26·0-33·4) and 13·1% (95%PI: 10·5-16·1) of all incident type-specific HPV detections could be attributable to transmission from a HITCH partner positive for that HPV type either at the start or end of the interval, respectively (Figure 1). Additionally, 14·5% (95%PI: 11·3-17·9) of incident type-specific HPV detections could be attributable to the participant reporting sex with other non-study partners during the interval. This leaves 42·7% (95%PI: 38·4-47·2) of incident HPV detections which could not be attributed to these risk factors for HPV transmission. The proportion non-attributable to sexual transmission was slightly higher in men (46·0%) than in women (39·0%).

**Figure 1.**
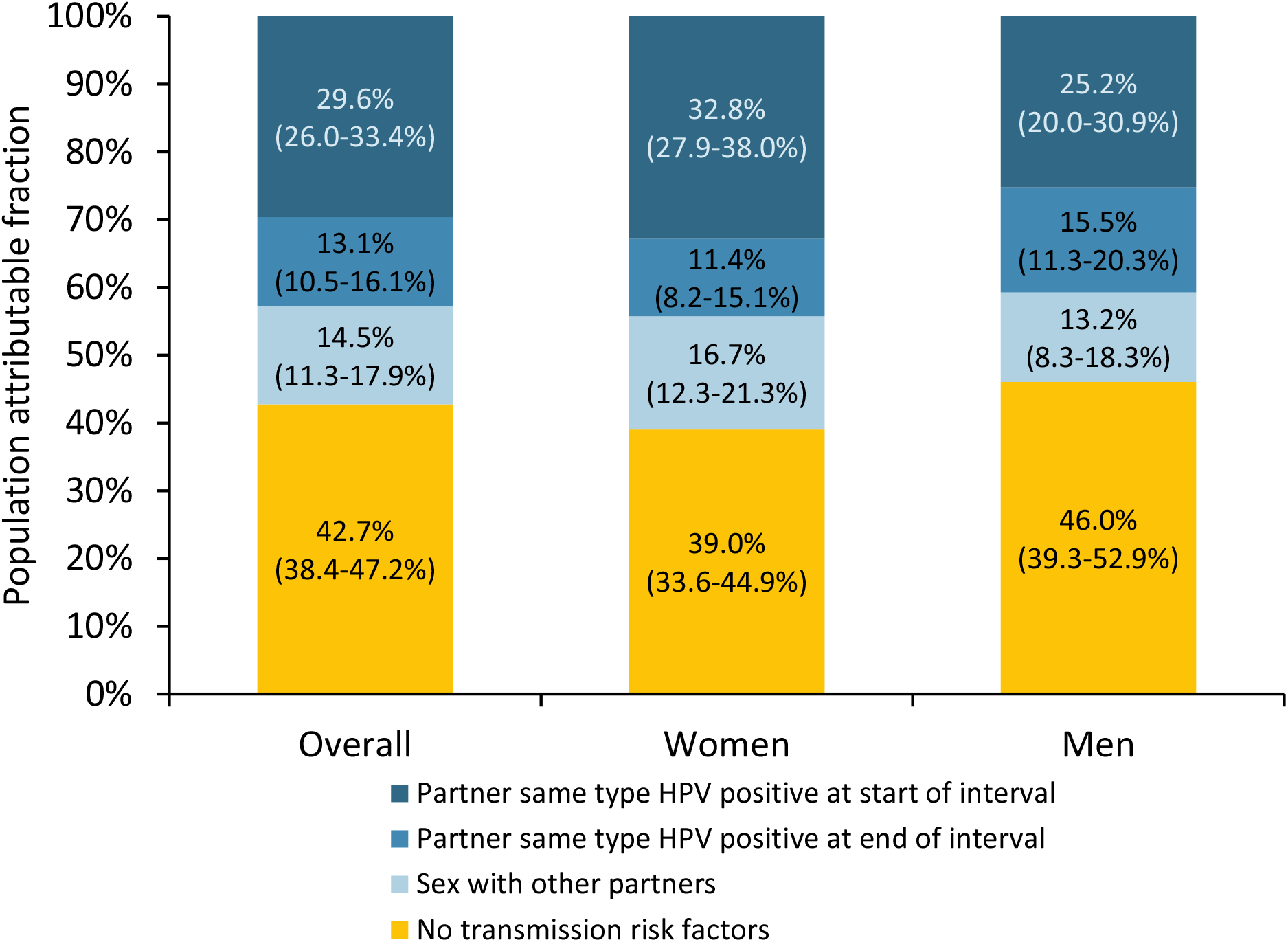
Population fraction of incident type-specific HPV detections attributable to sexual transmission risk factors, by sex. Numbers represent the median (2·5-95·5^th^ percentiles) of the posterior distribution. Dark blue: attributable fraction due to having a HITCH partner who is same-type HPV positive at the start of the interval. Medium blue: attributable fraction due to having a HITCH partner who is same-type HPV negative at the start but positive at the end of the interval. Light blue: attributable fraction due to having a HITCH partner who is same-type HPV negative at the start and negative/missing at the end of the interval, but reporting sex with other partners during the interval. Yellow: fraction of cases not attributable to any of the considered transmission risk factors.

To assess the impact of missing data for partner HPV status, we repeated the analysis considering missing partner HPV status as a separate category. Participants whose HITCH partner was type-specific HPV negative at the start of the interval and missing at the end of the interval, and who reported no other sex partners accounted for 72 (13·2%) incident type-specific HPV detections; they had a nearly identical incidence rate of HPV detection as participants whose HITCH partners were HPV negative for that type at both the start and end of the interval and reported no other sex partners (both 0·1/100 person-months), suggesting that combining these two categories was appropriate.

## Discussion

In this study of young heterosexual partners, although the majority (57%) of incident HPV detections could be attributed to sexual transmission, the remainder could not. These remaining 43% incident HPV detections are likely largely due to reactivation of latent or intermittent HPV infections.

An important limitation of this analysis was the high number of participants with missing values for their partner HPV status at the end of follow-up. It is possible we may have misclassified some of the incident HPV detections in these participants as being attributable to reactivation when they were instead due to transmission from a new HPV infection their partner acquired that we could not measure. However, because the incidence rate of detection was comparable to those with persistently HPV negative HITCH partners, and because many partners in this category had ended their sexual relationship, we believe that this misclassification is unlikely to be substantial. It is also possible some participants may not have reported having sex with other partners due to the sensitive nature of this question. This question, however, had a 99.9% response rate, with only one participant skipping the question. We also do not have data on which HPV infections participants had acquired prior to enrolment, and so cannot ascertain whether incident detections we assumed are reactivations were previously detectable in participants.

In a previous study of mid-adult women by Rositch *et al*.,^6^ 85% of incident HPV detections occurred in women who reported no new sexual partners, suggesting many incident HPV detections are likely attributable to reactivation rather than transmission. However, this study as well as others^4,7^ assessing HPV infections attributable to reactivation in women were individual-based studies. It is difficult with individual-level data to entirely exclude sexual transmission as a contributor to incident HPV detections, as it is not possible to control for partner HPV status. We recently performed a systematic review of couple-based studies of HPV transmission;^17^ to our knowledge the current study presents the first analysis assessing reactivation of HPV infections using a couple-based study design. While we found a lower proportion of infections that are potentially due to reactivation (43%) than Rositch *et al*., some of the difference between study estimates may be due to the younger age and higher level of sexual activity of our study population. The fraction of incident HPV detections attributable to reactivation is likely to be higher in less sexually active populations.

The risk of reactivation of an individual HPV infection is likely low; a previous study found redetection after HPV clearance occurred only in 7·7% of infections.^4^ However, the proportion of incident HPV detections due to reactivation in a population may still be high, because population attributable fractions are highly dependent on the prevalence of a risk factor.^14^ A low proportion of individuals are likely at risk of transmission due to having a new HPV positive sexual partner at a given point in time, while a large proportion of individuals may be at risk of reactivation of latent HPV infections from past exposures. We had previously found that HITCH participants who reported more lifetime sexual partners were more likely to have incident HPV detections than those with fewer lifetime sexual partner when their HITCH partner was HPV negative, as would be expected if these detections were reactivations from past exposures.^13^ Even though the incidence rate of type-specific HPV detection was very low in participants whose HITCH partner was currently negative for that HPV type, this group contributed the highest person-time at risk, and therefore accounted for a sizeable proportion of all incident HPV detections.

Finally, our results have two important implications for cervical cancer screening. Firstly, they suggest that women who have previously tested HPV-negative cannot be assumed to remain HPV-negative over time even if they have not had any new sexual partners. Multiple screenings over a lifetime are consequently important, despite the high long-term negative predictive value of a negative HPV test, due to both the risk of transmission and reactivation of latent HPV infections later in life. Secondly, our findings may help de-stigmatize a positive HPV screening test result; they suggest that a positive result in many cases is not indicative of recent sexual behaviors or partner infidelity, but rather a reactivated past infection.

## Data Availability

Participation consent forms specified that the participant data would only be published in aggregate form, and that individual-level participant data would only be made available to investigators involved in the HITCH study. To access individual-level HITCH data, please contact. The protocol for the HITCH cohort study has been published by El-Zein et al. 2019 (https://doi.org/10.2196/11284). Code used for the analysis can be found in the supplementary material.

## Contributions

ANB and ELF conceived and designed HITCH and obtained funding. ELF was the principal investigator for the study. ANB oversaw recruitment, data collection, provision of HPV test results to participants, and database design. PPT oversaw clinical activities and recruitment. FC supervised the laboratory analyses and the quality of PCR assays. MZ and ANB managed the HITCH database. TM and AM performed statistical analyses and wrote the first draft of the manuscript. ELF provided team supervision and advice. All authors read, provided feedback, and approved the final protocol and manuscript.

## Data sharing

Participation consent forms specified that the participant data would only be published in aggregate form, and that individual-level participant data would only be made available to investigators involved in the HITCH study. To access individual-level HITCH data, please contact Eduardo Franco at. The protocol for the HITCH cohort study has been published by El-Zein *et al*. 2019.^11^ Code used for the analysis can be found in the supplementary material.

