## Supplementary Material for "Proportion of new genital human papillomavirus detections attributable to latent infections: Implications for cervical cancer screening"

The code below was written in R 3.6.3 and requires the package R2WinBUGS.[1] The code implements a two-state Markov model to estimate transition rates based on the probability that an individual *i* will be HPV-positive (Infected[i]=1) or HPV-negative (Infected[i]=0) after a given follow-up time interval (intervalyears[i]), conditional on whether at the start of the interval they were HPV-positive (Infected_prev[i]=1) or HPV-negative (Infected_prev[i]=0) and given their category level for a risk factor of interest (category[i]). The model estimates the population attributable fractions (PAF[i] and PAFreflevel) for the risk factor of interest simultaneously using the estimated incidence rates for each level of the risk factor.

```
## Load R2WinBugs package
library(R2WinBUGS)

## Import dataset and format dataset for use by WinBugs as a list of variables
dataset<-read.csv(file="dataset.csv",header=T)
nobs<-nrow(dataset)
nID<-nlevels(factor(dataset$HITCHID))
varlist<-list(        nobs=nobs,
                      nID=nID,
                      HITCHID=dataset$HITCHID,
                      Infected_prev=dataset$Infected_prev,
                      Infected=dataset$Infected,
                      partnerInfected_prev=dataset$partnerInfected_prev,
                      intervalyears=dataset$intervalyears
                      nCategories=4,reflevel=1,
                      category=dataset$PAFcat,       ### Risk factor for PAF calculation
                      sumInfected=sum(dataset$Infected[dataset$Infected_prev==0]),
                      sumInfectedcat=colSums(sapply(1:4,function(x) dataset$Infected_prev==0 & dataset$Infected==1 &
                                   dataset$PAFcat==x)))

## Define a two-state model for HPV incident infections
Tmodel <-function() {

    #Assign random effects to each HITCHID
    #Each HITCHID has a random effect for the infections and clearance rates
    #A log-normal distribution random effects is assigned
    #lambda1 is the infection incidence rate
    #lambda2 is the clearance rate
    for (i in 1:nID) {
      loglambda1[i]~dnorm(meanL1,tau1)
      loglambda2[i]~dnorm(meanL2,tau2)
    }
    for (i in 1:nID) {
      log(lambda1[i])<-loglambda1[i]
      log(lambda2[i])<-loglambda2[i]
    }

    #Priors for rates and random effects
    meanL1~dnorm(0,0.1)
    meanL2~dnorm(0,0.1)
    sd1~dunif(0,4)
    sd2~dunif(0,4)
    tau1<-1/(sd1*sd1)
    tau2<-1/(sd2*sd2)

    #Priors for predictors of infection and clearance
    #beta2 is the ln(IRR) for clearance by partner infection status
    #beta1cat is the ln(IRR) for infection incidence rates by risk factor category
    #beta2cat is the ln(IRR) for clearance rates by risk factor category
    #alpha sets uniform prior weights for a Dirichlet for infection distribution by risk factor category
    #proportion represents the proportion of incident infections in each risk factor category
    beta2~dnorm(0,0.1)
    for (i in 1:(nCategories)) {
      beta1cat[i]~dnorm(0,0.1)
      beta2cat[i]~dnorm(0,0.1)
      alpha[i]<-1
    }
```

```
        proportion[1:nCategories]~ddirch(alpha[])
        sumInfectedcat[1:nCategories]~dmulti(proportion[],sumInfected)
        #Set IRR at reference level to 1
        beta1cat[reflevel]<-0
        beta2cat[reflevel]<-0

        for (i in 1:nobs){

          #Probability distribution of observed transitions
          Infected[i]~dbin(P[i],1)

          #Set up transition rates between HPV+ and HPV- states, with predictors
          #Incidence rate
          rate1[i]<-lambda1[HITCHID[i]] * exp(beta1cat[category[i]])
          #Clearance rate
          rate2[i]<-lambda2[HITCHID[i]] * exp(beta2cat[category[i]]) * exp(beta2*partnerInfected_prev[i])

          #Define transition probabilities between HPV+ and HPV- states in terms of rates
          #The following formula is the Kolmogorov solution for a two state model with backwards transitions

          P[i]<-(1-Infected_prev[i])*(1-((rate2[i]+rate1[i]*exp(-(rate1[i]+rate2[i])*intervalyears[i]))/(rate2[i]+rate1[i]))) +
                    Infected_prev[i]*((rate1[i]+rate2[i]*exp(-(rate1[i]+rate2[i])*intervalyears[i]))/(rate1[i]+rate2[i]))

        }

        #Parameters of epidemiological interest to calculate
        lambda1mean<-exp(meanL1)
        lambda2mean<-exp(meanL2)
        for (i in 1:nCategories) {
            IRcat[i]<-lambda1mean*exp(beta1cat[i])*10        #Incidence rates
            CRcat[i]<-lambda2mean*exp(beta2cat[i])*10        #Clearance rates
            IRR1cat[i]<-exp(beta1cat[i])                      #Incidence rate ratios
            IRR2cat[i]<-exp(beta2cat[i])                      #Clearance rate ratios

            #Population attributable fraction of incident infections due to risk factor
            PAF[i]<-(lambda1mean*exp(beta1cat[i])-lambda1mean*exp(beta1cat[reflevel]))/ (lambda1mean*exp(beta1cat[i]))*proportion[i]
        }
           #Population attributable fraction for the reference level
           PAFreflevel<-1-sum(PAF[1:nCategories])
}

## Names of parameters to output after simulation for further analysis:
parameters.PAF<-c("lambda1mean", "lambda2mean", "IRcat", "CRcat", "IRR1cat", "IRR2cat", "PAF", "PAFreflevel", "proportion")

## Create a function to set starting values of parameters for each simulation:
inits<-function(nID=nID, nCategories=4...){
      meanL1<-rnorm(1,-1,1); meanL2<-rnorm(1,-1,1)
      sd1<-1; sd2<-1;
      loglambda1<-rnorm(nID,meanL1,0.1)
      loglambda2<-rnorm(nID,meanL2,0.1)
      beta2<-0;
      beta1cat<-rep(0,nCategories); beta2cat<-rep(0,nCategories);
      proportion<-rep(1,nCategories)/nCategories;

      return(list(meanL1=meanL1, meanL2=meanL2, sd1=sd1, sd2=sd2, beta2=beta2, loglambda1=loglambda1, loglambda2=loglambda2,
                  beta1cat=beta1cat, beta2cat=beta2cat, proportion=proportion))
}

## Run model in WinBUGS:

bugs(model.file=Tmodel, data=varlist, parameters.to.save=parameters.PAF, inits=rep(list(inits()),3), n.chains=3, n.iter=50000,
n.burnin=20000, n.thin=5)
```
